# Longitudinal ctDNA Surveillance in Older Women with ER+ Breast Cancer to Facilitate Surgical De-Escalation: A Prospective, Hybrid-Decentralized Trial with Correlative Studies

**DOI:** 10.1101/2025.08.23.25332468

**Authors:** Neil Carleton, Alexander C Chang, Fangyuan Chen, Shannon L Puhalla, Julia Foldi, Hunter Waltermire, Antony Tin, Michael S Cowher, Kristin Lupinacci, Emilia J Diego, Quratulain Sabih, Ronald R Johnson, Monica Malhotra, Amanda Laubenthal, Vikram Gorantla, Marija Balic, Rohit Bhargava, Marion Joy, Tanner Freeman, Catherine Bridges, Ekaterina Kalashnikova, Angel Rodriguez, Minetta C Liu, Steffi Oesterreich, Adrian V Lee, Priscilla F McAuliffe

## Abstract

For older patients with competing comorbidities, optimizing oncologic therapies is of paramount importance. Circulating tumor DNA (ctDNA) is a validated prognostic factor across solid tumors and may provide a strategy to identify patients for whom safe de-escalation of certain therapies is possible. In this prospective, hybrid-decentralized trial (n = 43 patients; NCT05914792) that integrated clinical outcomes, patient- and caregiver-reported outcomes, and correlative tissue analysis, the primary objective was to determine if ctDNA levels were associated with tumor progression in older patients who opted to forgo breast cancer surgery in favor of primary endocrine therapy (pET). ctDNA levels were highly concordant with imaging findings, and a lack of ctDNA clearance at 6 months was associated with tumor progression. In a competing risk regression adjusted for patient age, tumor stage, tumor grade, and tumor Ki-67, pre-treatment ctDNA positivity was associated with a significant risk of tumor progression (HR 30, 95% CI 4.4-209; p = 0.0005). No patients with pre-treatment ctDNA negativity experienced tumor progression. In correlative analyses examining ctDNA-positive tumors progressing on pET, we identified populations of CD11+ T cell-interacting macrophages that upregulate CD109 and CD89 and secrete immunosuppressive chemokines to create a favorable environment for cancer epithelial cell proliferation. These findings suggest that ctDNA may be a surveillance modality for older patients who receive pET, warranting future evaluation in a randomized setting.

**STATEMENT OF SIGNIFICANCE:** Clinical management of older women with breast cancer can be challenging, and some women may opt to forego surgery through shared decision making with their physicians. Use of ctDNA for these patients may identify those at an increased risk for progression as well as those with endocrine-sensitive tumors who are good candidates for surgical de-escalation.

## Introduction

Clinical management of older patients (aged 70 years and older) with breast cancer can be challenging because tumor biology, life expectancy, competing risks of mortality, and patient preferences all play a role in determining optimal therapies (1-5). Recent pre-clinical and clinical evidence suggests that ER+ tumors arising in older patients tend to exhibit unique biologic underpinnings rooted in changes in estrogen disposition, chronic inflammation, and immune suppression, thus influencing their clinical presentation and behavior (6-9). This has led to recent changes in treatment paradigms that “right-size” therapy across the locoregional and adjuvant settings, largely through de-escalation or de-intensification (10-14).

Some older patients with ER+ breast cancer, either by their own choice or due to the recommendation of their treating physician, forgo locoregional intervention in favor of primary endocrine therapy (pET) (15, 16). In a retrospective analysis of 156 patients, we found that the risk of dying from breast cancer was 3% over 6 years when considering the risk of dying from another chronic condition (16). However, the rate of locoregional progression, where tumors presumably become resistant to endocrine monotherapy, remained elevated at 20%. While progression alone may not portend a poor outcome, as most patients respond to second-line endocrine monotherapy, locoregional disease may impair quality of life and ultimately warrant intervention if the patient is symptomatic (17). Critically, these data suggest that further optimization of non-surgical management for older women is warranted to identify patients most suitable for endocrine monotherapy or even active monitoring.

## Results

### Trial design and baseline clinical characteristics of enrolled patients

We designed a prospective longitudinal observational study for older patients who chose to forego surgery in favor of pET. The aim of the study was to assess if a personalized circulating tumor DNA (ctDNA) assay (Signatera^TM^ Exome, Natera) could identify patients at risk for progression on pET, as ctDNA offers both quantitative and longitudinal data to track on-treatment changes. Advantageously for older patients and their caregivers, having potential prognostic ability with minimally invasive testing facilitated by mobile phlebotomy may decrease healthcare contact days and the number of appointments. In this study, patients received ctDNA testing every 3-6 months at the same time as their standard-of-care imaging (most commonly, ultrasound of the breast and axilla) (**Fig. 1a**). Patients were monitored for response to and/or progression on pET. Patients could opt to stop pET at any point and undergo surgery, at which point their matched core biopsy and surgical specimens would be used for downstream correlative analysis. Forty-three patients were enrolled in the study, n = 34 of whom had a baseline / pre-treatment blood draw (CONSORT diagram in **Fig. 1b**). Median age at diagnosis was 86 years old (range: 75-94), and most patients had an ECOG Performance Status of 1 or 2, had tumors above 2cm in size without clinical nodal involvement, and received a non-steroidal aromatase inhibitor as their first-line pET modality (**Supplementary Table 1**; overall representativeness of the study population described in **Supplementary Table 2**). The median follow-up time was 2.1 years (range: 0.25-3.25 years). The study was designed in a decentralized manner: the majority of the patients (72.1%) were enrolled in community oncology clinics (**Supplementary Table 1**), and thus many of the patients relied on a caregiver (mostly adult children) to bring them to their appointments (**Fig. 1c**). Due to this, 70% of the patients opted for mobile phlebotomy to complete their longitudinal blood draws at home rather than returning to the oncology clinic (**Fig. 1d**).

**Figure 1:**
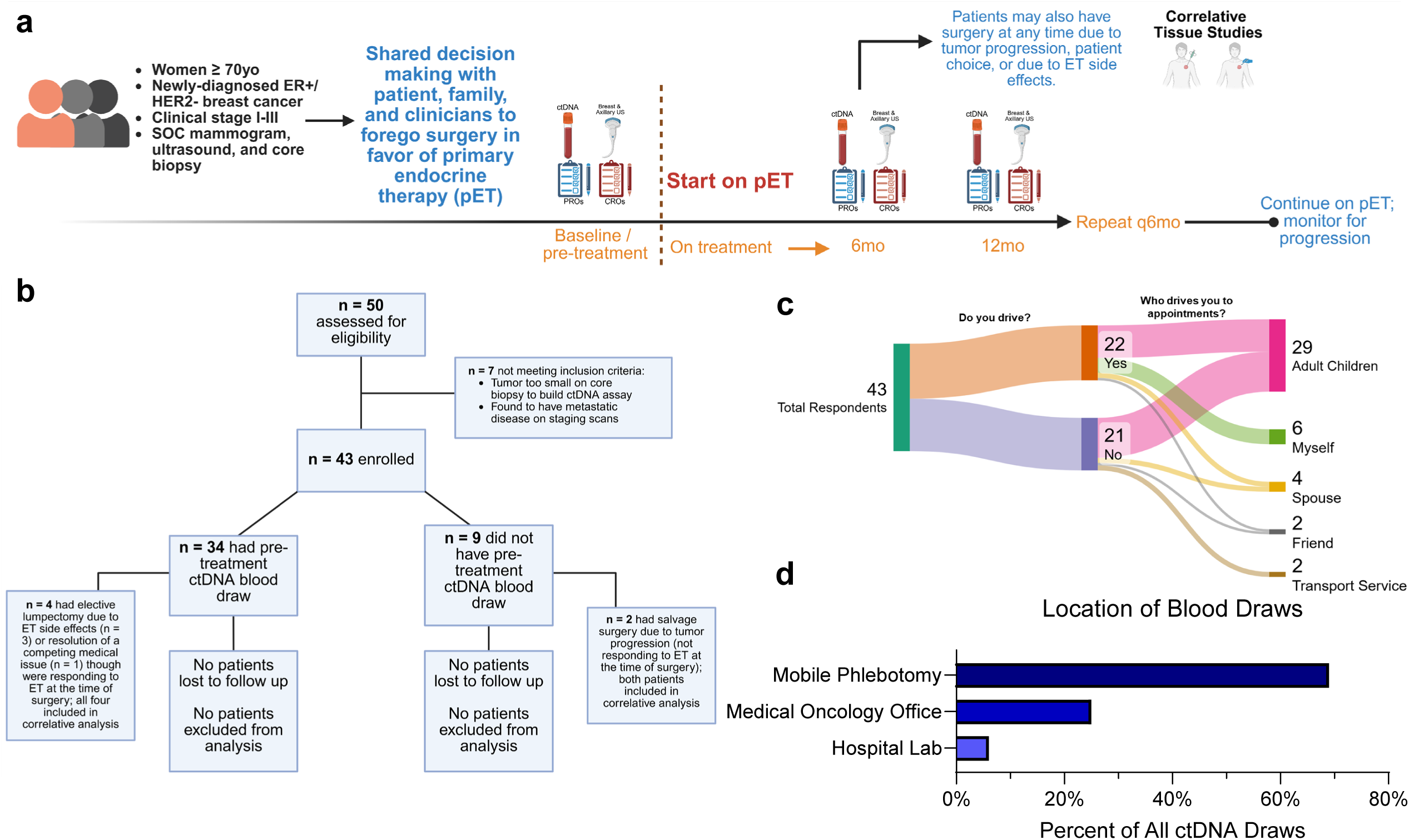
Overview of the study design and decentralized nature of the study. (a) Study schematic showing key inclusion criteria and study procedures. Patients could opt to receive surgery at any points, at which time their core biopsy and surgical specimens were procured for correlative studies. (b) CONSORT diagram for the study. (c) Sankey plot showing the driving habits of the 43 respondents demonstrated that decentralization of the study was essential, given that many of the patients in the study did not drive themselves to their appointments, instead relying on caregivers to do so. (d) Chart shows the location of where patients received their ctDNA blood draws. The majority of patients chose to receive their blood draws via Natera’s mobile phlebotomy service given that they did not drive any longer. Mobile phlebotomy service included going to patients’ homes, including nursing homes and assisted living facilities.

### ctDNA detection and association with clinical outcomes

Of those patients who had a pre-treatment ctDNA draw (n = 34), 32% had a ctDNA positive test result, with increasing frequency at higher levels of disease burden (**Fig. 2a**; additional swimmer plot of patients included in the study who did not have a baseline ctDNA draw can be found in **Supplementary Fig. 1**). Over the duration of the study, 5 out of 34 (15%) of patients experienced tumor progression; 1 out of 34 (2.9%) experienced death due to their breast cancer and 8 out of 34 (24%) died of non-breast cancer related causes. For patients who experienced progression, most opted for a change in systemic therapy, although palliative radiation and breast surgery were also performed. All clinical stage I (cT1N0) tumors were ctDNA negative at baseline, regardless of tumor grade or Ki-67 level, and remained negative with every subsequent test; none of these patients experienced progression.

**Figure 2:**
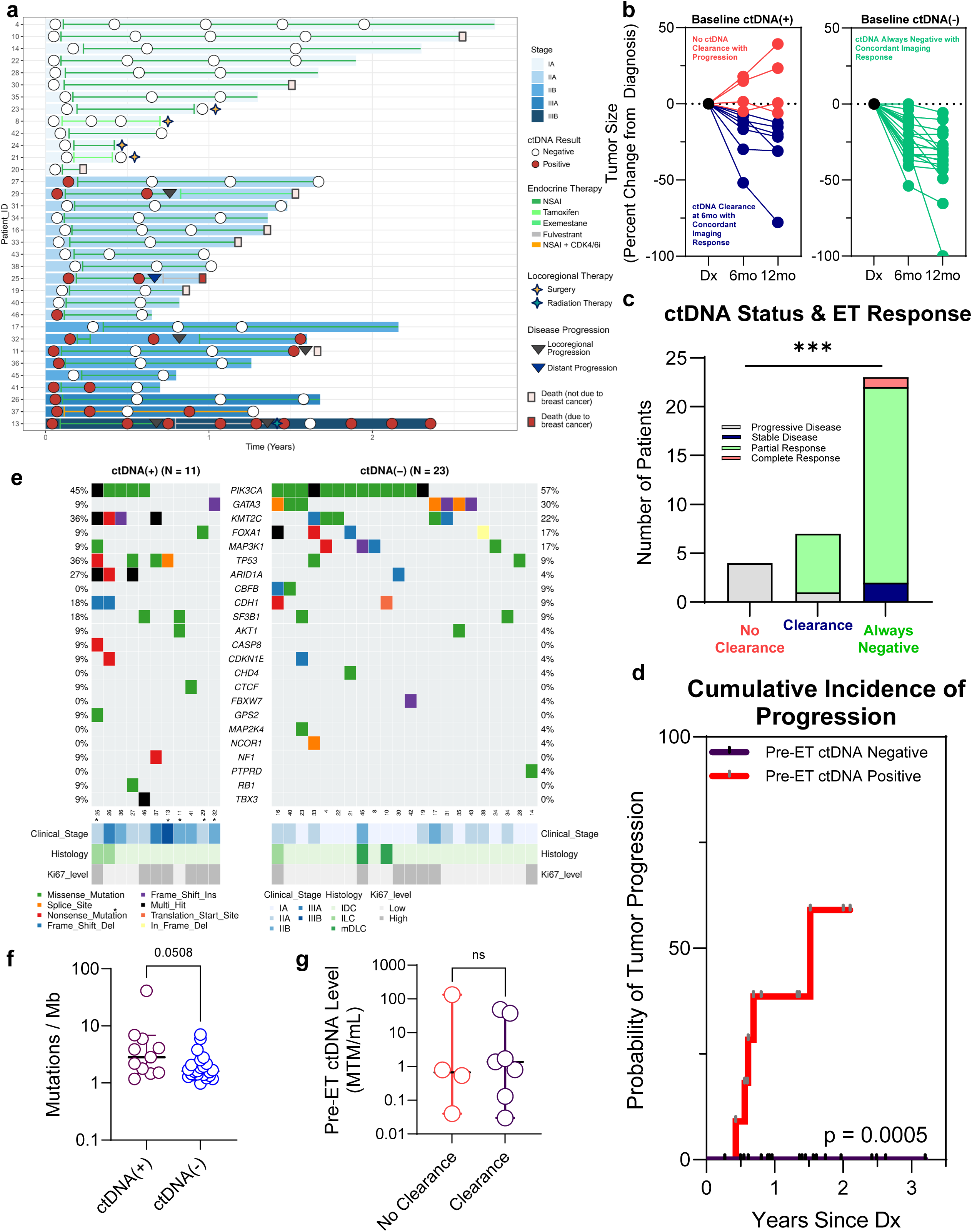
Longitudinal use of ctDNA identifies patients at high risk of progression and those who will durably respond to primary endocrine therapy. (a) Swimmer plot depicting ctDNA dynamics over time based on tumor stage at diagnosis. Overlayed onto the plot is the type of primary endocrine therapy (most patients started on a non-steroidal aromatase inhibitor (NSAI), such as anastrozole or letrozole), any type of locoregional intervention, disease progression, and vital status. (b) Plot shows the changes in tumor size based on ultrasound imaging based on the first 12 months on pET. Red indicates tumors that did not exhibit ctDNA clearance; blue indicates the tumors that were initially ctDNA positive at baseline but cleared ctDNA at 6 months; and green indicates the tumors that were ctDNA negative at baseline and remained ctDNA negative. (c) Bar plot showing association between ctDNA at 6 months with RECIST 1.1 outcomes. (d) Tumors that were ctDNA positive at baseline exhibited a significantly higher risk of progression using a competing risk regression analysis (p = 0.0005). (e) Using the core biopsies that were used for the ctDNA assay, we performed whole exome sequencing to identify potential genomic correlates of baseline (pre-treatment) ctDNA status. Within the baseline ctDNA positive group, those patients marked with an asterisk went on to experience tumor progression. (f) Though not reaching statistical significance, tumors that had baseline ctDNA positivity generally had higher levels of tumor mutational burden. (g) Within the baseline ctDNA positive group, there was no difference in baseline quantitative ctDNA level between those that cleared ctDNA at 6 months and those that did not clear ctDNA at 6 months.

ctDNA dynamics were highly concordant with changes in tumor size as assessed on serial breast ultrasound: tumors that exhibited baseline ctDNA positivity but cleared ctDNA by the 6-month draw, or had pre-treatment ctDNA negativity, all showed response to pET. Those that had pre-treatment ctDNA positivity without clearance at 6 months all had tumor progression (**Fig. 2b**). For one such patient on study who had over 10 draws on study, ctDNA was highly concordant across serial CT imaging and across multiple lines of endocrine therapy and palliative radiation therapy (**Supplementary Fig. 2**). When considering standardized responses to therapy using RECIST 1.1 criteria, 100% of patients without ctDNA clearance at 6 months experienced tumor progression. On the contrary, 100% of patients with baseline and persistent ctDNA negativity were without progression throughout follow up (p = 0.002) (**Fig. 2c**). Overall, in a competing risk regression adjusted for patient age, tumor stage, tumor grade, and tumor Ki-67, pre-treatment ctDNA positivity was associated with a significant risk of tumor progression (HR 30, 95% CI 4.4-209; p = 0.0005) (**Fig. 2d**).

We evaluated genomic correlates of ctDNA positivity from whole exome sequencing of the core biopsies. Tumors exhibiting ctDNA negativity were generally enriched for mutations associated with luminal, endocrine therapy-responsive disease, such as *PIK3CA*, *GATA3*, and *MAP3K1* (**Fig. 2e** showing common mutations found in breast cancer with an extended panel in **Supplementary Fig. 3**). Tumors exhibiting ctDNA positivity were associated with mutations that confer endocrine non-responsiveness, such as *TP53* and *ARID1A*, suggesting differences in underlying genomic biology between ctDNA positive and negative tumors. Concordantly, tumors with pre-treatment ctDNA positivity exhibited higher tumor mutational burden versus those that were ctDNA negative (p = 0.0508) (**Fig. 2f**). No underlying genomic differences were observed in tumors exhibiting ctDNA clearance versus those that exhibited persistent ctDNA positivity (**Fig. 2e**); the quantitative baseline ctDNA level did not differ between these groups (p = 0.79) (**Fig. 2g**).

### Patient- and caregiver-reported outcomes

Patients completed surveys every 6 months using the validated NFBSI-16 mechanism, and an additional accompanying survey had a series of study-specific questions (**Supplementary Material Display 1**). Over the course of the study, 93% of patients completed at least one survey. Overall, 82% of patients reported that they felt comfortable not undergoing surgery; however, 92% were at least “a little bit” concerned about the possibility of tumor growth or progression (**Fig. 3a**). Over 80% of patients felt that the results of the ctDNA blood draws could help inform their treatment plan. Patients reported tolerating pET well, with many patients indicating they felt only “a little bit” bothered by side effects (**Fig. 3b**). Patients had an 88.2% adherence rate to their pET over the course of the study, consistent with prior reports evaluating adherence in the pET setting (18). Patients rated the value of ctDNA highly throughout the study but felt it was most important in the first 6-12 months of testing (**Fig. 3c**). Importantly, patients did not experience anxiety related to the results of the ctDNA testing (**Fig. 3c**).

**Figure 3:**
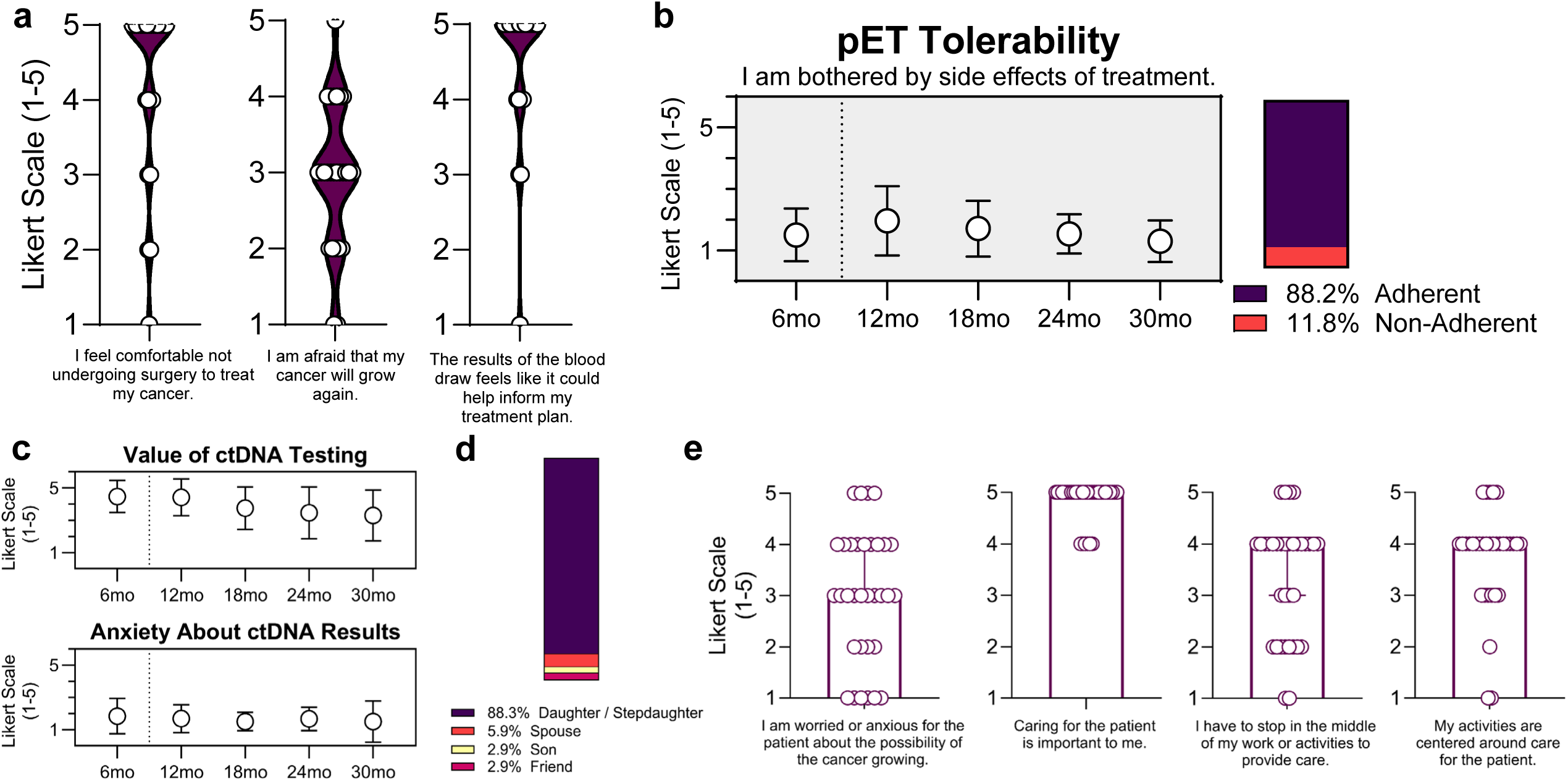
Patient- and caregiver-reported outcomes from the study showed that patients tolerated pET and did not have anxiety related to ctDNA testing and that caregivers had a high degree of caregiver burden. (a) Patient-reported outcomes showed, at baseline (pre-treatment), patients generally felt comfortable not undergoing surgery, felt that they were moderately afraid that the tumor would grow again, and felt that the results of the blood draws could help inform their treatment plan. Likert Scale (1-5): 1 = Not at all; 2 = A little bit; 3 = Somewhat; 4 = Quite a bit; 5 = Very much. (b) Over 88% of patients were adherent to their pET over the course of the study, with many of them reporting that they were not at all or only a little bothered by the side effects of therapy. Likert Scale (1-5): 1 = Not at all; 2 = A little bit; 3 = Somewhat; 4 = Quite a bit; 5 = Very much. (c) Patients highly valued ctDNA testing and generally had no or a little anxiety related to the results of ctDNA testing. (d) Caregiver reported outcomes showed that the vast majority of caregivers were the daughters, stepdaughters or daughters-in-law of patients in the study. (e) Caregivers were generally at least moderately worried about the possibility of the cancer growing, thought caring for the patient was very important, and had frequent interruptions in daily activities to take care of the patient. Likely Scale (1-5): 1 = Strongly Disagree; 2 = Disagree; 3 = Neither Agree nor Disagree; 4 = Agree; 5 = Strongly Agree.

Caregivers also completed surveys every 6 months using the Caregiver Reaction Assessment (19) with additional study-specific questions (**Supplementary Material Display 2**). The majority of patients had caregivers present during their visits (86%), and of those caregivers, 89% completed at least one survey. We observed that the vast majority of caregivers attending visits for patients on study were the adult daughters, stepdaughters, or daughters-in-law (**Fig. 3d**). The majority of these caregivers were “at least somewhat” worried about the possibility of tumor growth, which did not change over time (**Fig. 3e**). While caregivers indicated that caring for the patient was very important, most noted that they had to stop or reduce working and other activities (within the past 3-6 months) to care for the patient, indicating a degree of caregiver burden.

### Multifaceted tumor microenvironment (TME) changes underlie ctDNA-positive progression on long-term endocrine monotherapy

Biospecimens were collected for 6 patients who underwent breast surgery while on trial, creating a unique cohort of matched core biopsy and surgical specimens after long-term endocrine monotherapy. Four patients’ tumors were responding to pET at the time of surgery but they decided to stop pET due to side effects (n = 3) or resolution of competing medical issue (n = 1), while two patients had tumor progression at the time of surgery (**Fig. 4a**; clinical characteristics of these patients in **Supplementary Table 3**). All patients responding to pET exhibited longitudinal ctDNA negativity, while both patients experiencing locoregional progression showed positive ctDNA. The two patients with progression both had initial partial responses to pET with eventual progression after 36-48 months on therapy (**Supplementary Fig. 4a**). At the time of diagnosis, there were no major underlying genomic differences in the six tumors (**Supplementary Fig. 4b**).

**Figure 4:**
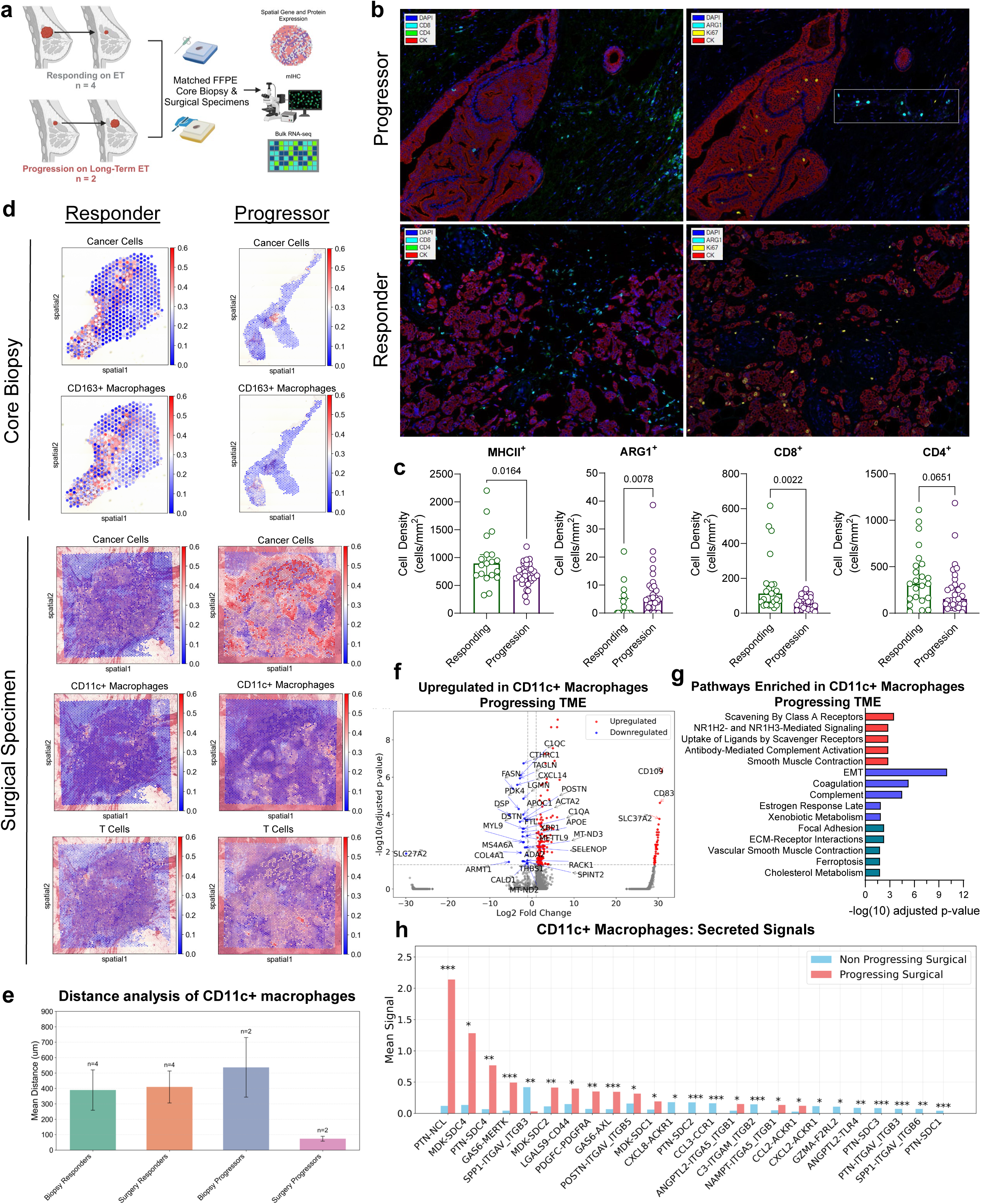
Spatial transcriptomic analysis of long-term endocrine monotherapy tumors indicates a reversion to an immunosuppressive TME in progressing samples. (a) Schematic of correlative studies. (b) Multiplex immunohistochemistry of FFPE blocks from the surgical specimens (responding to pET and progressing on pET) examining stromal T cells (CD4 and CD8) and macrophages showed decreased infiltration of T cells and higher levels of ARG1+ macrophages in the progressing TME. White box highlights a population of stromal ARG1+ macrophages in the progressing TME. (c) Quantification from the multiplex immunohistochemistry from stromal regions of the TME. (d) Spatial plots showing cancer epithelial cells and CD163+ macrophages in core biopsies and cancer epithelial cells, CD11c+ macrophages, and T cells in the surgical specimens. (d) Distance analysis of the macrophages relative to the cancer epithelial cells from the spatial plots showing that in the progressing specimens, the macrophages were in closer proximity to the progressing, proliferating cancer epithelial cells. (f) Volcano plot showing differential gene expression analysis of the macrophages in the progressing vs. responding TME shows upregulation of CD109 and CD89, both linked to immunosuppressive TGF-β signaling. (g) Pathway enrichment in the CD11c+ macrophages in the progressing TME shows activation of classically pro-tumor scavenging receptors. Pathways in red indicate those from KEGG; blue are from REACTOME; and purple are from HALLMARK. (h) CD11c+ macrophages in the progressing TME showed secretion of factors such as CCL2, pleiotrophin (PTN), midkine (MDK), and LGALS9; taken together, these factors have been classically linked to exclusion and exhaustion of infiltrating T cells. * p < 0.05, ** p < 0.01, *** p < 0.001.

We utilized spatial transcriptomic sequencing to provide a deeper interrogation of the on-therapy TME, comparing those responding (ctDNA negative) at the time of surgery versus those progressing (ctDNA positive). Focusing on the cancer epithelial cells, the responding cancer epithelial cells showed upregulation of ECM pathways, RAGE signaling with DAMP creation in the TME, and protein digestion (**Supplementary Fig. 6a**). The progressing cancer epithelial cells were enriched for pathways related to MYC, oxidative phosphorylation, and mTORC signaling (**Supplementary Fig. 6b**). Though genomic information was not available on the tumors that showed progression, we found an ESR1 mutation (D538G) in one of the surgical specimens (**Supplementary Fig. 6c**), which likely also contributed to pET resistance.

Using multiplex immunohistochemistry to quantify the local immune infiltrate, we observed that there were significantly fewer CD8+ T cells, CD4+ T cells, and MHCII+ macrophages and significantly more ARG1+ macrophages in the progressing TME compared to the responding TME (**Fig. 4b**, **Fig. 4c, and Supplementary Fig. 5**), suggesting a role for immunosuppressive macrophages in the progressing TME. To evaluate this hypothesis, we further characterized macrophages identified in the progressing and responding TME.

The progressing TME showed an expected proliferation of cancer epithelial cells with a colocalized CD11c+ macrophage infiltrate (**Fig. 4d**). On distance analysis, macrophages in the progressing specimens were closer to the cancer epithelial cells than those found in the core biopsies and the responders (**Fig. 4e**). Macrophages in the progressing TME showed upregulation of genes such as *CD109* and *CD89* (**Fig. 4f**) and enrichment of pathways related to scavenger receptors and EMT activity (**Fig. 4g**). Conversely, macrophages found in the responding TME showed activation of innate immune response with ROS production (**Supplementary Fig. 6d**). When we evaluated secreted signals from the macrophages into the TME, the macrophages in the progressing TME showed significantly higher levels of pleiotrophin, midkine, CCL2, and LGALS9 secretion compared to the macrophages in the responding TME (**Fig. 4h**). Collectively, these data suggest that the CD11c+ macrophages in the progressing specimens revert to an immunosuppressive, pro-tumor phenotype. Overall, after pET, the responding (ctDNA negative) TME showed a marked decrease in cancer cell burden, a stromal macrophage infiltrate characterized by CD11c+ macrophages, and a prominent T cell infiltrate.

## Discussion

An important goal for treatment of older patients with breast cancer is “right-sizing” therapy: adequately treating the breast cancer to avoid locoregional and distant complications while balancing quality of life and minimizing potential overtreatment and exposure to drug toxicity. In this prospective, observational hybrid-decentralized trial, we evaluated longitudinal ctDNA assessment to monitor treatment escalation and/or de-escalation in patients who choose to forgo locoregional therapy in favor of pET. We found that baseline (pre-treatment) ctDNA positivity conferred a risk for tumor progression. Risk was highest among patients who had persistent ctDNA positivity after six months of pET. ctDNA dynamics were highly concordant with changes in tumor size as assessed via serial breast ultrasound. Patients acknowledged that ctDNA testing gave them additional information to better understand their tumor behavior.

A particular strength of the study is its patient-centered nature. Many older patients receive their care in community oncology sites, and a vast majority of the patients in this study were enrolled from such locations. Because the study offered mobile phlebotomy services, patients could participate in serial blood draws from their own homes, including assisted-living sites. As trials become more decentralized (20), offering convenient options for trial participation, particularly for older individuals, who often do not drive themselves to appointments (including serial imaging, surgical oncology, and medical oncology), could alleviate some of the caregiver burden identified in this study. ctDNA testing in particular may offer additional data about a tumor’s response to therapy without having patients return to the office for as many interval imaging studies and appointments.

The association between ctDNA dynamics and clinical outcomes in the study was notable. All of the patients with stage I disease had no detectable ctDNA. This may either be due to the sensitivity of the ctDNA assay or due to the often-indolent tumor biology of small, luminal tumors in older patients. ctDNA negativity was highly associated with favorable responses to pET. After 6 months of pET, a persistent ctDNA positive result was strongly associated with increase in tumor size on ultrasound, whereas persistent ctDNA negative result or clearance of ctDNA at 6 months was associated with tumor stability or size reduction on ultrasound. The majority of the progressions occurred locoregionally with either growth of the primary tumor or axillary lymph nodes. This 6-month window may offer a potential time-point for intervention for those that do not clear the ctDNA. Those patients with persistently positive ctDNA may in fact be poor candidates for pET and they may need additional intervention to achieve tumor control. This may be completed through locoregional therapy or through additional systemic therapy options; in this study, most patients opted for a change in systemic therapy though palliative radiation was also well-tolerated. In this study, we found that salvage fulvestrant was logistically challenging, given the need to return to the office monthly for injections. Oral selective estrogen receptor degraders are a promising option for second-line pET in this setting. Interestingly, the quantitative baseline ctDNA level of those that cleared ctDNA at 6 months, and those that did not, was not significantly different.

These findings suggest an opportunity to refine patient selection for treatment escalation or de-escalation in future trials. For example, ctDNA positivity either at baseline or persisting after a defined treatment interval (i.e., 6 months) could serve as a biomarker-enriched eligibility criterion for escalation arms in prospective studies. In contrast, patients with sustained ctDNA negativity may be ideal candidates for de-escalated surveillance strategies without additional treatment intensification. Incorporating ctDNA dynamics as a stratification factor in such trials may help personalize therapy based on real-time tumor biology rather than static clinicopathologic features alone. Ultimately, this could contribute to a framework for older adults in whom both over and under treatment carry significant risks.

An additional strength of this study was the embedded hypothesis-generating correlative results. On-therapy biospecimens from patients treated with long-term endocrine monotherapy are rare, and few groups have studied the immunologic changes in those responding and progressing on pET. While we did identify one progressor with an *ESR1* mutation, there were shared immune responses between both progressing patients. We mainly focused on the tumor-associated macrophages in the TME, which have a well-documented role for supporting the development and growth of ER+ breast tumors (21, 22). Interestingly, we observed an eradication of CD163+ immunosuppressive macrophages in the responding TME. During progression, however, there seemed to be a re-emergence of an immunosuppressive, pro-tumor state marked by CD11c+/ARG1+ macrophages. Sequencing of events leading to tumor progression remain murky, but it is likely that a combination of changing tumor genomic features, local immunologic alterations, and accumulation of higher burdens of systemic chronic inflammation are all contributory (23). Indeed, though it is recognized that cancer cell-extrinsic mechanisms support endocrine therapy resistance and cancer cell persistence, there has been little direct evidence linking macrophages to this phenomenon to date (24, 25). Prior studies have shown that mitochondrial bioenergetics and epigenetic alterations may also assist in nongenetic mechanisms of cancer cell persistence. Recent evidence suggests that ER+ tumors which persist after neoadjuvant endocrine therapy undergo mitochondrial reprogramming and shuffle ATP production through oxidative phosphorylation (26), which was one of the pathways we also identified in this study in the tumors which progressed on pET. Though unexplored in this study, stochastic epigenetic reprogramming has also been shown to facilitate cancer cell persistence in response to endocrine therapy (27). Though most of this work has mainly been studied in vitro and in the setting of reawakening dormant cells in metastatic disease, similar mechanisms may underlie tumor persistence in those patients who are treated with long-term endocrine monotherapy.

This study has several limitations. Most importantly, both the ctDNA analysis and the correlative science aspect of the study included small samples sizes. Given the hypothesis-generating nature of the study, future randomized studies utilizing ctDNA monitoring to risk-stratify patients are warranted. This study also had a relatively short follow up time; however, older patients with significant multimorbidity often have numerous competing causes of death that preclude long-term follow up. In this study, only one patient died due to their breast cancer – many others passed from other causes, such as congestive heart failure, kidney disease, and infection. Nevertheless, this study provides compelling pilot data that ctDNA may be a means to identify patients who may safely de-escalate certain therapies in the treatment of their breast cancer and may be a robust surveillance modality for those receiving pET.

## Methods

### Clinical trial design

This was a prospective, single-center, observational (non-randomized) study, utilizing a clinically-validated personalized, tumor-informed multiplex PCR-NGS assay (Signatera™, Natera, Inc.) to monitor changes in circulating tumor DNA (ctDNA) in older (≥70 years) women with early-stage, estrogen receptor-positive (ER+) breast cancer who elect to forgo surgical management in favor of pET. Inclusion criteria required participants aged 70 and older with histologically confirmed ER+/HER2- non-metastatic (stage I-III) breast cancer who decided to forgo surgery and who initiated pET. Trial enrollment required adequate tumor tissue available for ctDNA assay development. Exclusion criteria were evidence of metastatic disease (staging scans were performed as clinically indicated at the discretion of the treating physician), prior systemic therapy for a different cancer, and inability to provide serial blood samples. There was no blinding in the study, and both patients and physicians were informed of the serial ctDNA and imaging results. Full study demographics can be found in **Supplementary Table 1**. Patients (n = 43) were recruited from April 2022 through April 2024.

The trial was registered at clinicaltrials.gov under NCT05914792. The Institutional Review Board at the University of Pittsburgh gave ethical approval for this work (STUDY 21100091). The study was conducted in accordance with the Declaration of Helsinki (as revised in 2013). Informed consents were obtained from all participants.

### Study objectives, endpoints, and procedures

The primary objective was to identify rates of ctDNA detection in women ≥ 70 years old with stage I-III breast cancer, the correlation of ctDNA result with clinical progression, and the feasibility of ctDNA monitoring as a tool to guide management decisions. Secondary endpoints included cumulative risk of tumor progression and patient- and caregiver-reported outcomes.

Participants in the trial signed consent forms and underwent their first draw on study within 2-4 weeks. Patients then underwent longitudinal ctDNA blood draws every 3-6 months at the discretion of their treating physician.

Corresponding with the timing of the ctDNA draws, patients completed patient-reported outcome surveys and underwent standard-of-care breast and axillary ultrasound, with optional mammogram. Patient-reported outcomes were assessed using the NCCN NFBSI-16 survey mechanism with an additional set of study specific questions (a copy of the survey can be found in **Supplementary Material Display 1**). Patient-reported outcome surveys could either be completed in the office during patients’ surgical and medical oncologist follow up appointments or could be completed via mailed out copies with pre-paid return envelopes. Caregiver-reported outcomes were assessed using the Caregiver Reaction Assessment with an additional set of study specific questions (a copy of the survey can be found in **Supplementary Material Display 2**). Caregiver-reported outcome surveys were completed during follow up appointments.

Patients could also undergo CT imaging at the discretion of their treating physician. Patients could either receive their longitudinal blood draws via Natera’s mobile phlebotomy service or at the time of their oncology appointments. Follow up for progression occurred until the time of death. Patients could opt to undergo surgery at any point. Tumor progression was based on RECIST 1.1 criteria. For patients who opted to stop ctDNA testing, their electronic health record was checked periodically for progression and vital status. No patients were lost to follow up or excluded from analysis.

### ctDNA assay and whole exome sequencing

Circulating tumor DNA (ctDNA) was assessed using the Signatera™ molecular residual disease (MRD) assay (Natera, Inc., Austin, TX), a personalized, tumor-informed multiplex PCR next-generation sequencing (mPCR-NGS) platform. For each participant, a tumor tissue sample and a matched normal blood sample was used to identify up to 16 somatic, single-nucleotide variants (SNVs) through whole-exome sequencing. Following this, a personalized mPCR-NGS assay was developed for each individual to longitudinally quantify ctDNA from plasma. Blood samples were collected in Streck tubes and processed for cell-free DNA extraction according to the manufacturer’s protocol. The patient-specific ctDNA assay was tested on associated cell-free DNA libraries via mPCR, and subsequent sequencing was performed on an NGS platform. Plasma samples with >2 SNVs detected above a predefined algorithm confidence threshold were considered ctDNA-positive. ctDNA concentrations are reported as mean tumor molecules per milliliter of plasma (MTM/mL).

### Correlative studies

Patients who underwent surgery during the study had their archived specimens procured for correlative studies, including spatial transcriptomics, bulk RNA sequencing, and multiplex immunohistochemistry.

### 10X Visium library preparation and sequencing

Before sectioning, formalin fixed paraffin embedded (FFPE) samples were assessed for RNA quality, and all samples had a DV200 score above the minimum threshold (≥30%). Spatial transcriptomic and proteomic libraries were generated with Visium CytAssist for the FFPE v2 gene expression kit (10x Genomics: 1000520) with the Human Immune Cell Profiling Panel (10x Genomics: PN-1000607).

Five-micron FFPE sections were placed on Schott Nexterion Hydrogel Coated Slides (Schott North America: 1800434) or Fisherbrand Superfrost Plus Slides (Fisher: 22-037-246) and processed through the 10x CytAssist for FFPE v2 protocol according to the manufacturer’s instructions. Library QC was completed with an Agilent TapeStation 4150. Libraries were normalized and pooled to 2 nM before loading on an Illumina NextSeq 2000 using a P3 100 flow cell. The pooled library was loaded at 650 pM, and sequencing was carried out with a 28/10/10/50 base pair read structure, targeting 125 million reads per sample for transcriptomic libraries, and 25 million reads per sample for proteomic libraries.

### Total RNA library generation and sequencing

RNA was extracted from FFPE sections using the Purelink FFPE RNA isolation kit (Invitrogen: K156002). According to the manufacturer’s instructions, RNA-seq libraries were generated with the Takara SMART-Seq Stranded kit (Takara: 634447). RNA was normalized to 5 ng/μl in a total volume of 7 μl of input RNA. RNA fragmentation was not performed.

Ten cycles were used for PCR1, followed by depletion of ribosomal RNA using scZapR, and 12 cycles were completed for PCR2. Library quantification and evaluation were performed using a Qubit FLEX fluorometer and an Agilent TapeStation 4150. Libraries were normalized and pooled to 2 nM before sequencing on an Illumina NextSeq 2000 using a P4 200 flow cell. The pooled library was loaded at 750 pM. Sequencing was done with a 2×101 bp read structure, targeting 40 million reads per sample. A custom run chemistry was used to incorporate three dark cycles at the start of R2 to mask the three bases of the Takara adapter present in the read. Sequencing data was demultiplexed using the onboard Illumina DRAGEN FASTQ Generation software.

### Spatial transcriptomic data analysis

CITEgeist methodology is described in detail elsewhere (28). In brief, CITEgeist is implemented in Python 3.10 using Gurobi 11.0.2 for ILP optimization. Data preprocessing includes gene filtering (> 0 counts in ≥ 1% spots, mean expression > 1.1) and normalization (target sum 10,000). Antibody data undergoes winsorization for the top and bottom 5% of datapoints and global centered log ratio normalization. COMMOT (29), GSEAPY (30), and PyDeSeq2 (31) were used as described in their respective vignettes. COMMOT was run with a 1% cutoff for signals screened. GSEAPY gene lists were derived from Scanpy. Wilcoxon Rank Gene Group results with an adjusted p-value less than 0.05. PyDeSeq2 was run with size factors fit type set to ‘poscounts’ due to the sparse nature of spatial data.

For cell type-specific transcriptional analysis, a cell type-specific differential gene expression analysis was conducted to uncover transcriptional changes within individual cell populations across different conditions. For each cell type identified by CITEGeist, we aggregated its estimated gene expression profiles from all spots across all samples where its proportion exceeded 0.2. This created a new, cell-type-specific AnnData object. To correct for inter-sample technical variability, we applied Harmony batch correction to these aggregated profiles, using the sample of origin as the batch key. Subsequent analysis on the corrected data included dimensionality reduction via PCA and UMAP, followed by Leiden clustering. Differential gene expression analysis was performed using the Wilcoxon rank-sum test (scanpy.tl.rank_genes_groups) to identify transcriptional differences between conditions (biopsy vs. surgical) and clinical outcomes (progressing vs. non-progressing) within each specific cell type.

To perform an integrated analysis of all cell states across all samples, we constructed a unified dataset. For each spatial spot, the CITEgeist-estimated gene expression profiles for all constituent cell types were expanded to create a comprehensive AnnData object where each observation represents a unique cell state in a specific spatial location. This integrated object, which combines data from all biopsy and surgical samples, was then batch-corrected using Harmony to mitigate technical artifacts between samples. On this integrated and corrected dataset, we performed principal component analysis, UMAP for visualization, and Leiden clustering to identify shared cell-state communities across all conditions. Finally, we conducted differential gene expression analysis using the Wilcoxon rank-sum test to compare transcriptional programs between experimental conditions (biopsy vs. surgical) and clinical outcomes (progressing vs. non-progressing) at a global, integrated level.

Intercellular communication was inferred using the COMMOT framework to model signaling networks within the tumor microenvironment. For each sample, we utilized the CITEGeist-derived cell-type-specific gene expression profiles and their corresponding spatial coordinates to model ligand-receptor interactions and downstream pathway activities. The resulting sender and receiver signaling scores for hundreds of pathways were aggregated at the cell-type level. To identify signaling dynamics associated with treatment response and progression, we performed statistical comparisons of pathway activity scores between our defined sample cohorts (non-progressing vs. progressing and biopsy vs. surgical). Significantly altered pathways were identified and visualized using bar plots and split-signal plots to dissect the contributions of sender and receiver cells to the differential signaling observed between conditions.

### Multispectral immunohistochemistry

Automated multiplex staining of FFPE tissue sections was performed on the Leica Bond RX. For staining, Akoya Bioscience’s Opal 6-Plex Detection Kit (cat# NEL871001KT) was used according to the manufacturer’s instructions for sequential staining of each marker in each panel. Akoya Bioscience’s PhenoimagerHT 2.0 platform and InForm® 3.0 analysis software were used for whole slide scanning, spectral unmixing, and cellular phenotyping.

### Statistical analysis and reporting summary

Descriptive statistics, chi-squares testing, and independent sample t-testing were used to analyze baseline clinical characteristics. Competing risk regression to evaluate the cumulative risk of tumor progression was analyzed using the Fine-Gray method. Tumor progression was defined as the time from diagnosis to the time of tumor growth meeting RECIST 1.1 criteria.

P-values were calculated using a two-sided method. Statistical significance was defined by a p-value less than 0.05. GraphPad Prism (RRID: SCR_002798) and Stata (Version 17; College Station, TX, USA) were used for statistical calculations.

## RESOURCE AVAILABILITY

### Materials Availability

Materials and reagents can be requested from the lead contact upon reasonable request. This paper did not generate unique reagents.

### Data and Code Availability

All raw and processed data from the correlative studies are available through GEO (accession GSE289326; token for reviewers: ohylkmmyjpcbrmr). All analysis code is publicly released in a GitHub repository (https://github.com/leeoesterreich/ctDNAManuscript) to ensure transparency and reproducibility.

## ACKNOWLEDGEMENTS

We thank all patients who consented for the study and their caregivers. We also thank the UPMC Hillman Cancer Center’s Clinical Research Services for their support of this study. This project used the

Hillman Cancer Center Translational Pathology Imaging Laboratory that is supported in part by award P30CA047904 and the NSABP Foundation.

Work performed in the UPMC Hillman Cancer Center Tissue and Research Pathology/Pitt Biospecimen Core and services and instruments used in this project were supported, in part, by the University of Pittsburgh, the Office of the Senior Vice Chancellor for Health Sciences, and the UPMC Hillman Cancer Center under award P30CA047904. This project utilized the services of the University of Pittsburgh Health Sciences Sequencing Core at UPMC Children’s Hospital of Pittsburgh. This study used high performance research computing core (RRID: SCR_022735) at the University of Pittsburgh Center for Research Computing supported by NIH S10OD028483 (to AVL).

This project was funded through the Hillman Cancer Center Developmental Pilot Program (to SO and AVL), the Shear Family Foundation (to SO and AVL), and the National Institutes of Health under awards 5T32CA82084-20 (to NC), 5F30CA264963-03 (to NC), P30CA047904, and S10OD028483 (to AVL).

## Author Contributions

Conceptualization: AVL, PFM, NC, SO, CB, EK, AR, MCL

Methodology: NC, ACC, FC, AT, RB, MJ, TF, CB, EK, AR, MCL, SO, AVL, PFM

Investigation: NC, ACC, FC, HW, MSC, KL, EJD, JF, SLP, QS, RRJ, MM, AL, VG, MB, SO, AVL, PFM

Visualization: NC, ACC, FC, MJ, AT, AVL, PFM

Funding Acquisition: AVL, SO, NC

Project Administration: PFM, AVL, SO, NC

Supervision: PFM, AVL, SO, AR, MCL

Writing – Original Draft: NC, PFM, AVL, ACC

Writing – Review & Editing: All authors.

## Declaration of Generative AI and AI-Assisted Technologies

The authors did not use generative AI or AI-assisted technologies in any aspect of the writing of this manuscript.

## Supplemental Information

- Supplementary Material: contains Supplementary Table 1 through 3 and Supplementary Figures 1 through 6.
- Supplementary Display 1 and 2 are appended with copies of the patient- and caregiver-reported outcome surveys.

