## Supplementary Material for "Longitudinal ctDNA Surveillance in Older Women with ER+ Breast Cancer to Facilitate Surgical De-Escalation: A Prospective, Hybrid-Decentralized Trial with Correlative Studies"

### Supplementary Table 1

| | | All patients (total n = 43) |
|---|---|---|
| **Age at Enrollment, median (range)** | | 86 (75-94) |
| **Risk Analysis Index (RAI)# at Enrollment, median (range)** | | 36 (35-44) |
| **ECOG Performance Status (%)** | | |
| | 0 | 4 (9.3%) |
| | 1 | 21 (48.8%) |
| | 2 | 15 (34.9%) |
| | 3 | 3 (7.0%) |
| **Patient Race / Ethnicity (%)** | | |
| | Non-Hispanic White | 37 (86.0%) |
| | Black | 3 (7.0%) |
| | Asian | 3 (7.0%) |
| **Clinical T Stage (%)** | | |
| | T1 | 15 (34.9%) |
| | T2 | 21 (48.8%) |
| | T3-T4 | 7 (16.3%) |
| **Clinical N Stage (%)** | | |
| | N0 | 31 (72.1%) |
| | N1-N2 | 12 (27.9%) |
| **Palpable Tumor on Physical Exam (%)** | | |
| | Yes | 30 (69.8%) |
| | No | 13 (30.2%) |
| **Tumor Histology (%)** | | |
| | IDC / NST | 37 (86.0%) |
| | ILC | 4 (9.3%) |
| | mDLC | 2 (4.7%) |
| **Tumor Ki-67 (%)** | | |
| | Low (< 20%) | 19 (44.2%) |
| | High (≥ 20%) | 24 (50.0%) |
| **Enrollment Location (%)** | | |
| | Acacdemic | 12 (27.9%) |
| | Community Clinic | 31 (72.1%) |
| **Pre-Treatment ctDNA Draw (%)** | | |
| | Yes | 34 (79.1%) |
| | No | 9 (20.9%) |
| **Staging Scan Prior to Starting ET (%)** | | |
| | Yes | 6 (13.9%) |
| | No | 5 (11.6%) |
| | Not Clinically Indicated | 32 (74.4%) |
| **Type of ET (%)** | | |
| | Anastrozole | 34 (79.1%) |
| | Letrozole | 6 (14.0%) |
| | Tamoxifen | 2 (4.7%) |
| | AI + CDK4/6i* | 1 (2.2%) |

**Supplementary Table 1:** Baseline clinicopathologic characteristics of patients enrolled in the study.
Abbreviations: IDC / NST = invasive ductal carcinoma or carcinoma of no special type; ILC = invasive lobular carcinoma; mDLC = mixed ductal-lobular carcinoma; ET = endocrine therapy.
### RAI calculated based on patients with cancer, which would give a minimum score of 35. RAI can be calculated manually at: https://efrailty.hsl.harvard.edu/ToolRiskAnalysisIndex.html (PMID: 27893030).
* For this patient, they started on anastrozole with ribociclib.

### Supplementary Table 2

| Characteristic | Cohort in current study (NCT05914792); n = 43 | US reference (SEER) of older patients diagnosed with new breast cancer (restricted to the those diagnosed ≥ 75 years) |
|---|---|---|
| Median age (range) | 86 (75–94) | In the US, patients aged 75-84 comprise 14.6% of all new breast cancer diagnoses; those over the age of 85 comprise 4.9% of all new diagnoses. |
| Stage at diagnosis | T1: 34.9%; T2: 48.8%; N0: 72.1% | Localized ~70%; Regional ~24%; Distant ~6.5% |
| Histology | IDC/NST: 86%; ILC: 9.3%; Mixed: 4.7% | NST ~75%; Lobular ~10–15% |
| Subtype ($HR^+/HER2^-$) | All $ER^+$ (majority $HR^+/HER2^-$) | ~70% $HR^+/HER2^-$ |
| Race/Ethnicity | 86% White, 7% Black, 7% Asian | ~60–70% White; ~10–15% Black; ~5–10% Asian; ~10% Hispanic |
| Geography / Site type | 28% Academic; 72% Community | ~70–80% of older adults treated in community settings |
| Other Considerations | All participants ≥ 75 y (median RAI 36) with 42% ECOG ≥ 2, reflecting a frail population. Trial included only patients managed with endocrine monotherapy. Cohort was 86% White with no Hispanic participants. 72% of participants enrolled from community clinics. | These considerations support generalizability to frail, older $ER^+$ breast cancer patients, but healthier or more diverse populations are underrepresented. |

**Overall Representativeness of the Study**

The study cohort is broadly representative of very elderly (≥ 75 years), frail patients with $ER^+$ breast cancer in the United States. The high proportion of community-based enrollment, distribution of clinical stage, and histology align with SEER data, although the cohort underrepresents Hispanic patients and healthier older adults who may receive more aggressive therapy.

This study cohort (n = 43; all ≥ 75 years, median age 86, median RAI 36, and 42% ECOG ≥ 2) closely mirrors the demographic and clinical features of prior primary endocrine therapy (pET) cohorts in older women with $ER^+$ breast cancer. Carleton et al. (PMID 38135542) reported a median age of 82 (range up to 97), with ~82% $ER^+/PR^+$ disease, 85% T1–T2 tumors, and 66% node-negative, which closely parallels our distribution of 83% T1–T2 and 72% N0. Similarly, large observational pET studies (PMIDs 37479944, 36326972) describe patient populations with median ages of 81-83, predominantly frail, with early-stage, unifocal, $ER^+$ tumors and low nodal burden. In these series, 70-85% of patients were node-negative, most tumors were histologic grade 1–2, and selection for pET was strongly associated with comorbidity or functional dependence – characteristics directly reflected in our cohort with high RAI scores and ECOG ≥ 2. Collectively, our study population is highly representative of the real-world population of elderly, frail women receiving pET for $ER^+$ breast cancer in contemporary practice, supporting external validity for this therapeutic context.

### Supplementary Fig. 1

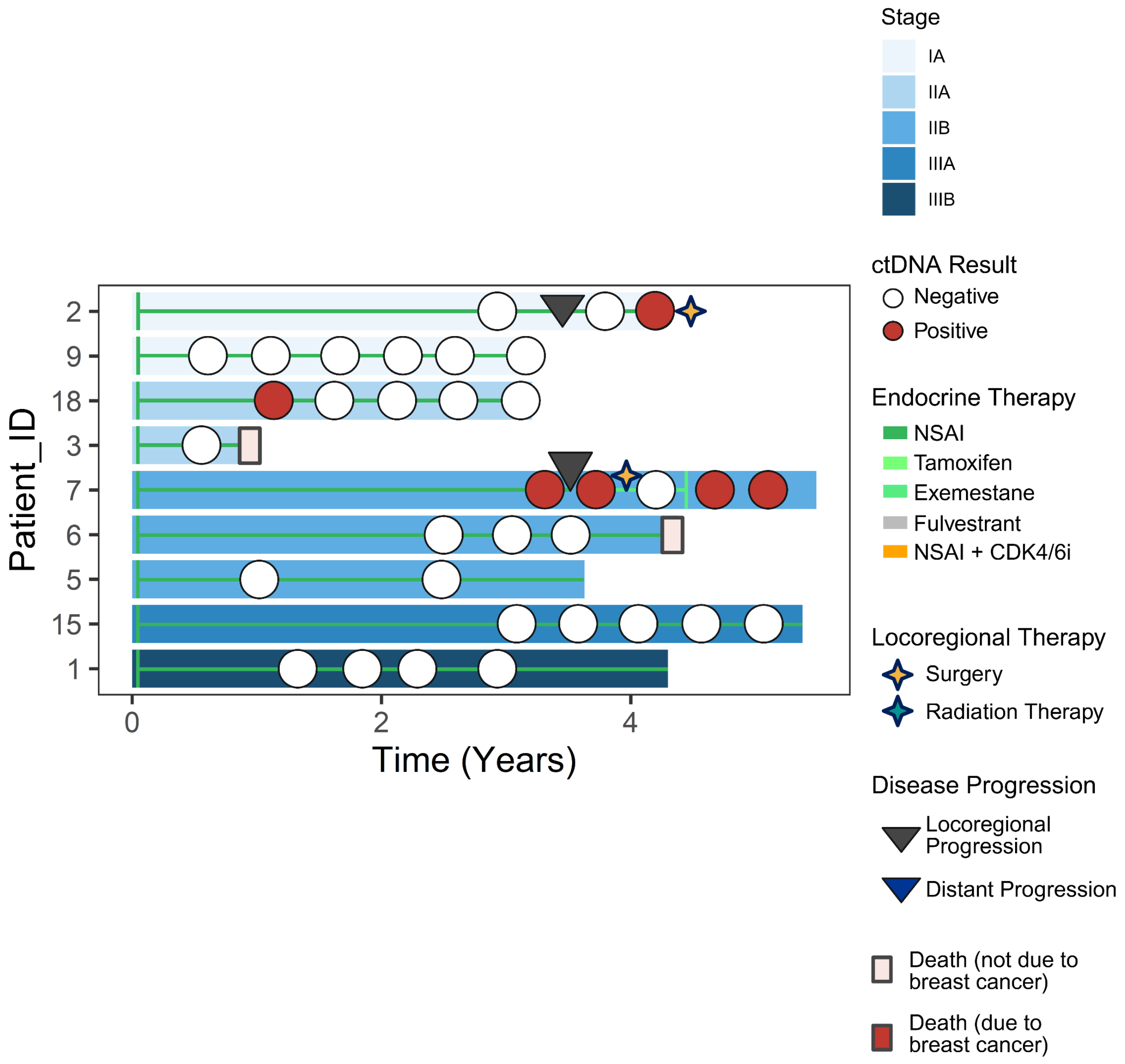


**Supplementary Figure 1:** Swimmer plot for those patients without a baseline (pre-treatment) ctDNA draw but were still enrolled in the study. Overlayed onto the plot is the type of primary endocrine therapy (most patients started on a non-steroidal aromatase inhibitor (NSAI), such as anastrozole or letrozole), any type of locoregional intervention, disease progression, and vital status.

### Supplementary Fig. 2

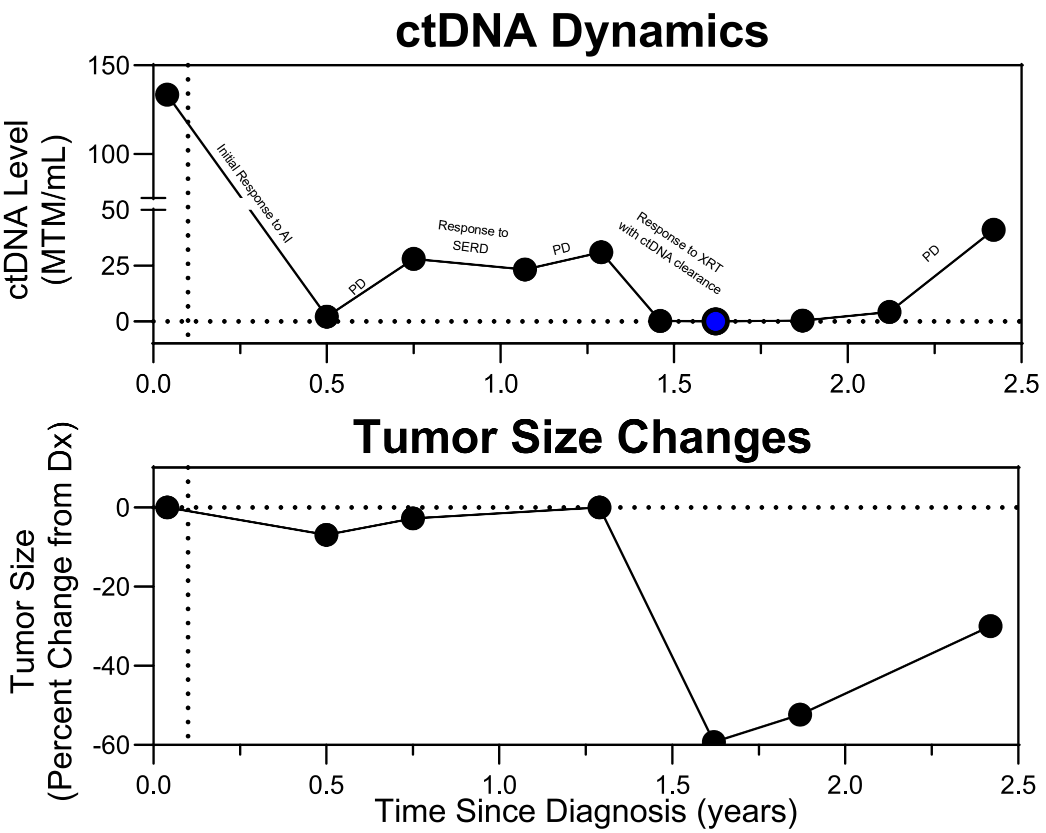


**Supplementary Figure 2:** ctDNA dynamics of one patient (Study ID: 13) enrolled on the study who had 10 draws over the course of the study period. This patient experienced multiple progressions (as indicated in the plot) with intermittent durable responses to endocrine therapy and palliative radiation therapy. Accompanying the ctDNA dynamics plot is the tumor size changes based on volumetric tumor size on CT scans.

### Supplementary Fig. 3

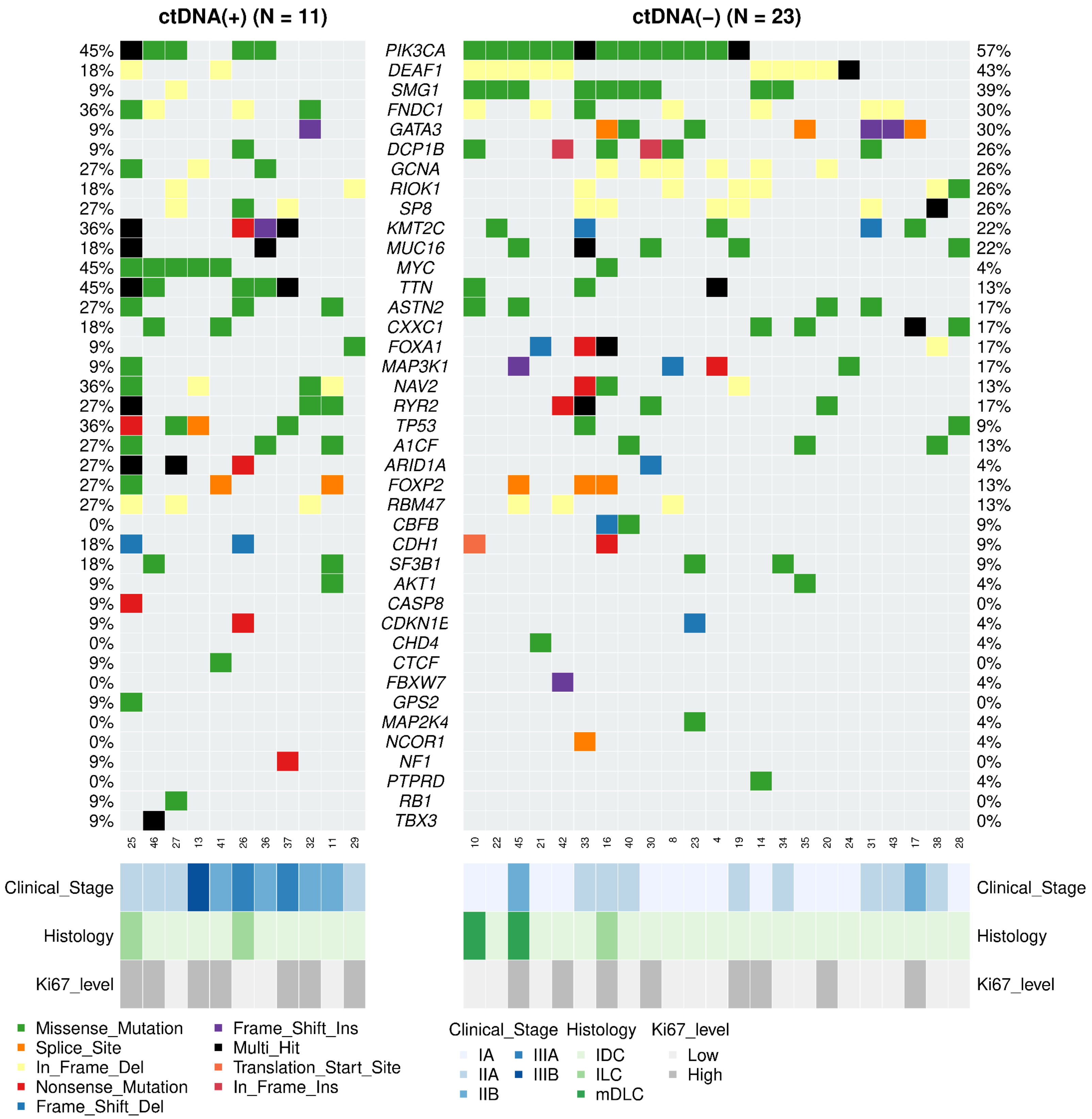


**Supplementary Figure 3:** Whole exome sequencing from the core biopsies of all patients with a baseline (pre-treatment) ctDNA blood draw. Plot shows the top 20 mutated genes in addition to those commonly found in breast cancer.

### Supplementary Table 3

| Patient_ID | Age Range at Dx | Tumor Histology | Clinical Tumor Stage at Dx | Months on ET | Response to ET | ctDNA Status at Surgery |
|---|---|---|---|---|---|---|
| 8 | 76-80 | IDC | cT1bN0 | 9 | Partial Response | Negative |
| 21 | 81-85 | IDC | cT1bN0 | 6 | Partial Response | Negative |
| 24 | 76-80 | IDC | cT1bN0 | 6 | Partial Response | Negative |
| 23 | 76-80 | IDC | cT1bN0 | 13 | Partial Response | Negative |
| 2 | 86-90 | IDC | cT1cN0 | 48 | Partial Response then Progressive Disease | Positive |
| 7 | 76-80 | ILC | cT2N0 | 42 | Partial Response then Progressive Disease | Positive |

**Supplementary Table 3:** Clinicopathologic characteristics of patients included in the correlative studies. All four patients who were responding to pET at the time of surgery had baseline ctDNA negativity. Of those with progressing tumors, one patient had baseline ctDNA negativity and was ctDNA negative at the time of progression and surgery (patient 2); one patient did not have ctDNA testing at baseline but was ctDNA positive at the tie of progression and surgery (patient 7).
Abbreviations: IDC / NST = invasive ductal carcinoma or carcinoma of no special type; ILC = invasive lobular carcinoma; mDLC = mixed ductal-lobular carcinoma; ET = endocrine therapy.

### Supplementary Fig. 4

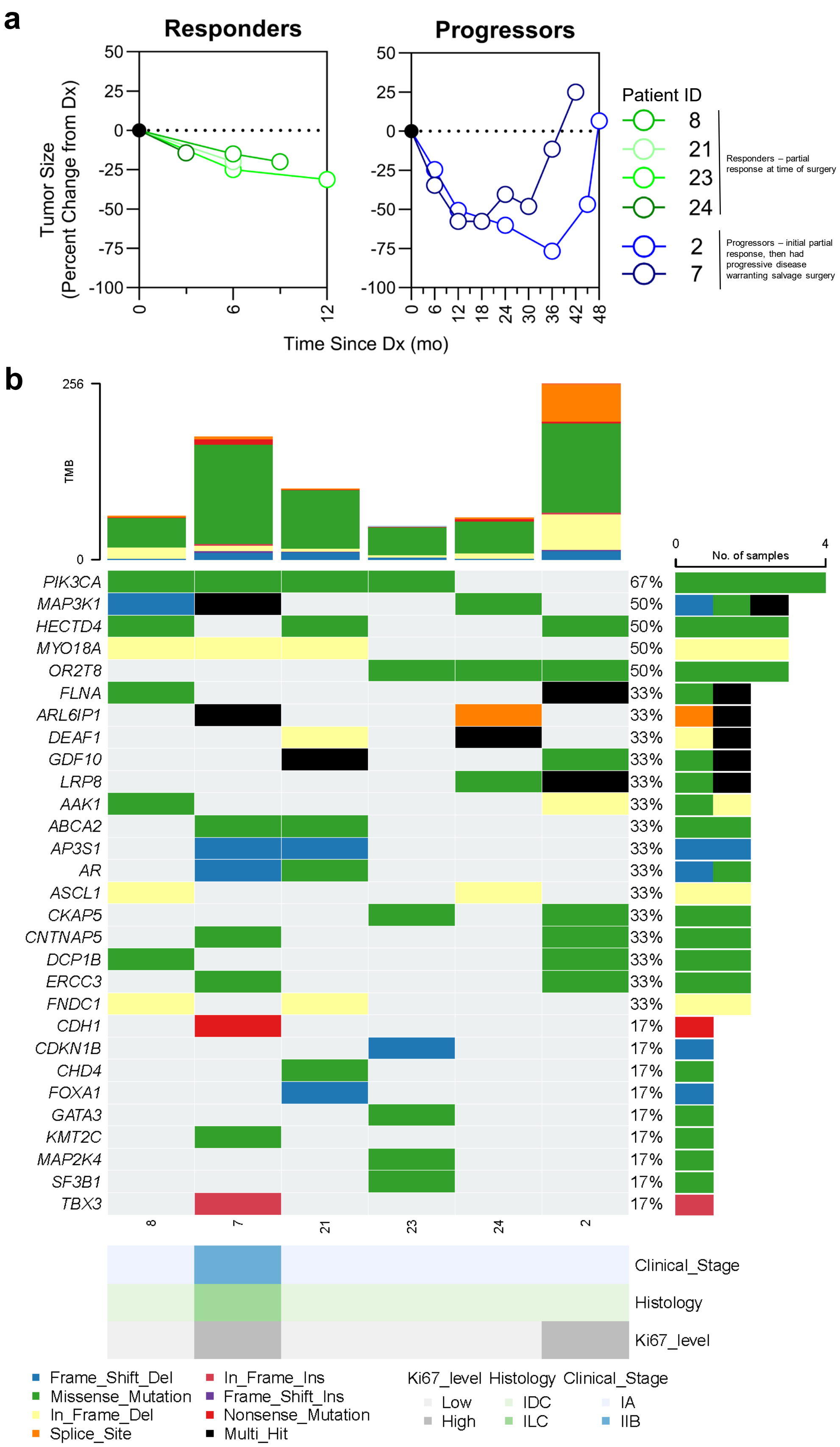


**Supplementary Figure 4:** (a) n = 6 patients, with matched core biopsy and surgical specimens, were included in the correlative studies: n = 4 patients had tumors that were responding to pET at the time of surgery while n = 2 patients had tumors that were progressing at the time of surgery. Notably, the two patients who experienced tumor progression did initially have a partial response. (b) Whole exome sequencing from the core biopsies of these six patients at the time of their initial diagnosis shows no underlying genomic differences. Plot shows the top 20 mutated genes in addition to those commonly found in breast cancer.

### Supplementary Fig. 5

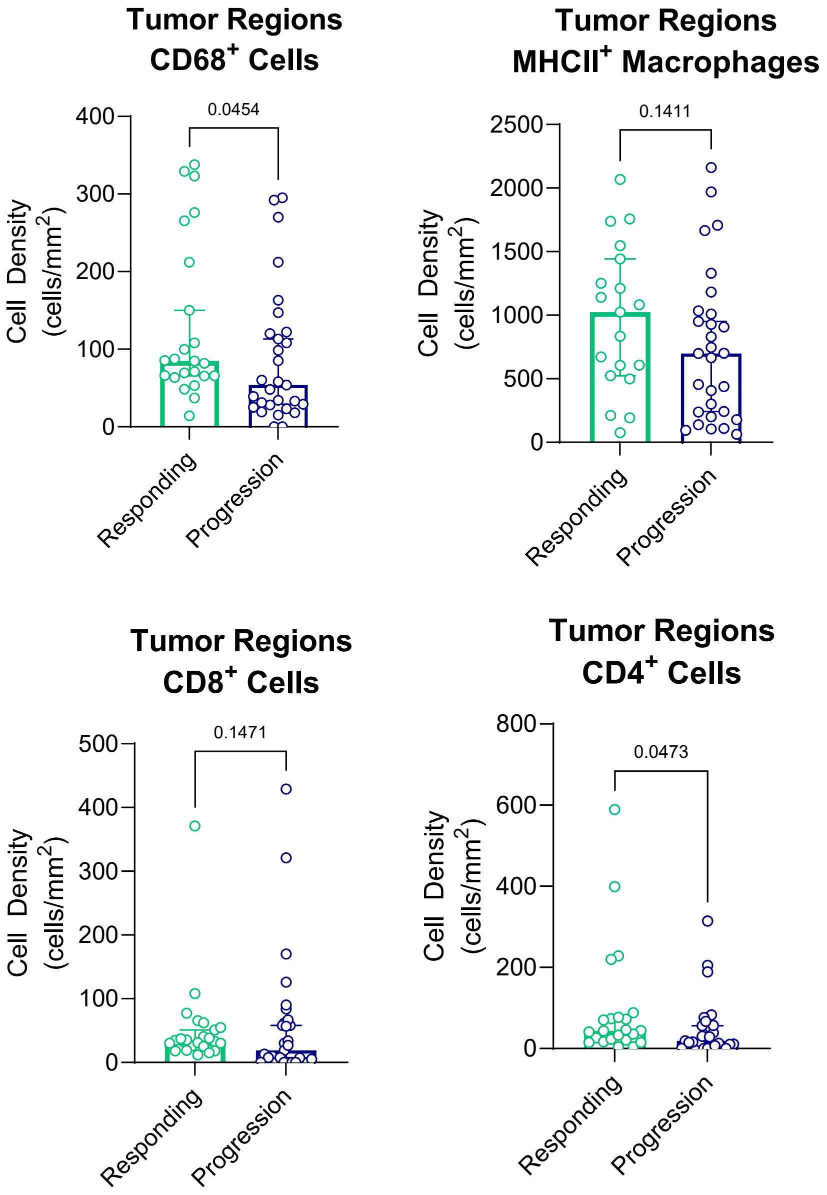


**Supplementary Figure 5:** Quantification of T cells and macrophages from the multiplex immunohistochemistry in stromal regions of the TME.

### Supplementary Fig. 6

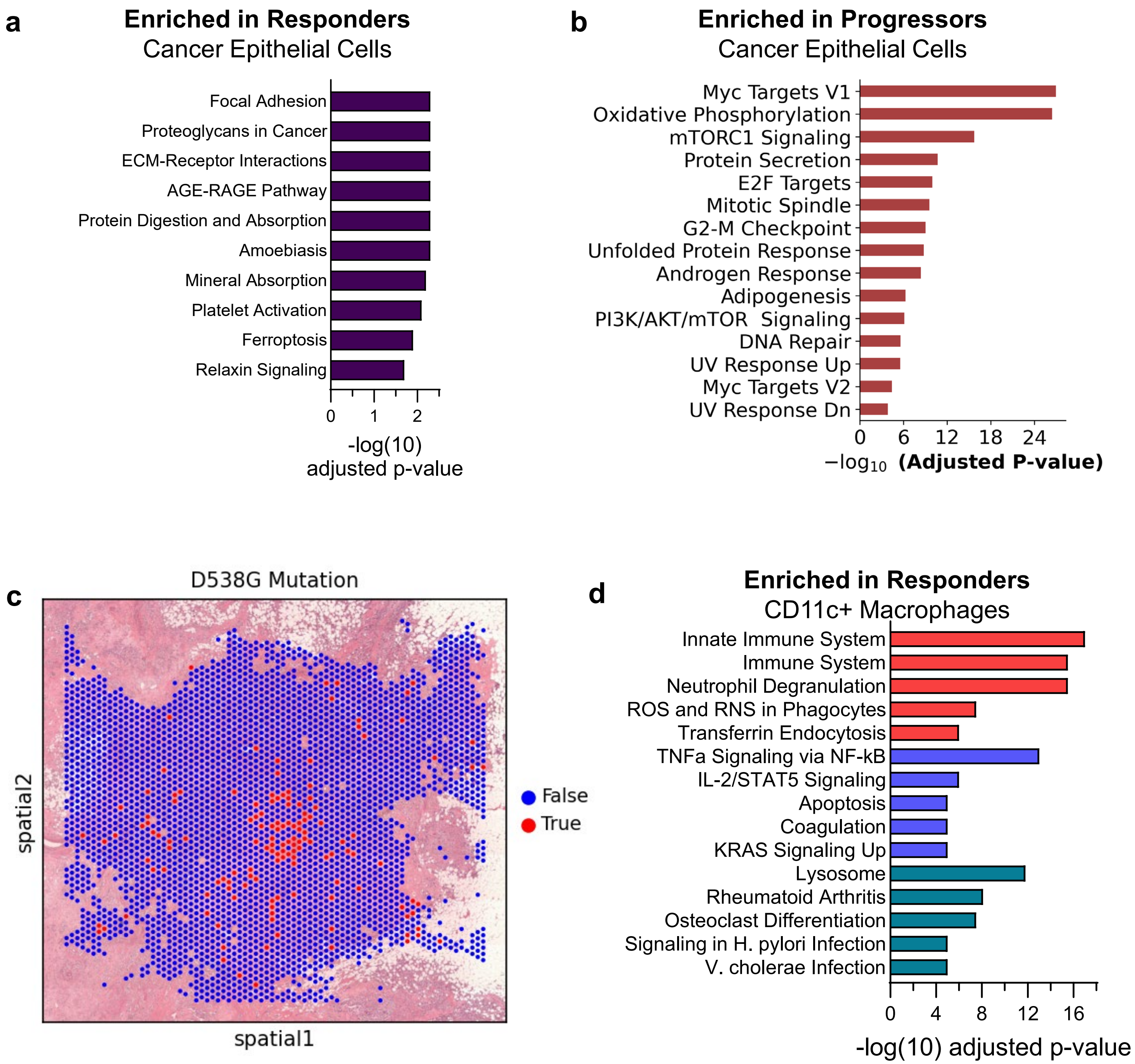


**Supplementary Figure 6:** (a) Pathway enrichment from the cancer epithelial cells in the responding TME. (b) Pathway enrichment of cancer epithelial cells in the progressing TME. (c) Detection of cancer cells harboring an ESR1 mutation in one patient progressing on pET. Both patients progressing on pET were tested. The other patient did not have an ESR1 mutations. (d) Pathways enriched in macrophages in the responding TME. Pathways in red indicate those from KEGG; blue are from REACTOME; and purple are from HALLMARK.
